# *Lactobacillus pentosus* KCA1 Decreases Vaginal and Gut Microbiota Associated with Bacterial Vaginosis (BV), Down-regulates IL-1 beta in Women of Child-Bearing Age and Modulates Bacterial Genes Related to Metabolic Functions

**DOI:** 10.1101/2020.04.02.20040238

**Authors:** Kingsley C Anukam, Chinwe E Ejike, Nneka R Agbakoba, Charlotte B Oguejiofor

## Abstract

**Introduction:** Bacterial vaginosis which affects 14-50% of reproductive-aged women in Nigeria is misdiagnosed and under-reported. Treatment option is antibiotics, which leads to recurrent infections. The objectives of this study are three folds, first to determine effects of oral feeding of *Lactobacillus pentosus* KCA1 on the vaginal and gut microbiota of women diagnosed with BV; to measure the level of two proinflammatory cytokines IL-1 beta, before and after KCA1 consumption and to determine the relative abundance of bacterial metabolic genes.

**Methods:** Seven women diagnosed with BV by Nugent score (7-10) were recruited to provide vaginal and gut sample before and after 14 days oral intake of 3 grams of *Lactobacillus pentosus* KCA1. The DNA from the swabs were processed for 16S rRNA metagenomics using Illumina MiSeq platform. The paired-end sequence FASTQ reads were imported into Illumina Basespace pipeline for quality check (QC). In addition, EzBioCloud pipeline was use for alpha and beta diversity estimation using PKSSU4.0 version and open reference UCLUST_MC2 for OTUs picking at 97% cut-off. Blood samples were analyzed using ELISA technique. PICRUSt was used to predict the metabolic functions from the 16S rRNA gene dataset.

**Results:** On average, there was no significant difference at p=0.05 in the alpha indices typified by Shannon index. The beta diversity showed different clustering positions with PCoA. However, at individual taxonomic categories, there was a significant decrease in the relative abundance of some genera associated with bacterial vaginosis after KCA1 feeding with a corresponding increase of *Lactobacillus* genera. Bacterial genes related to defence systems were up-regulated in the vagina. There was a 2-fold down-regulation of IL-1 beta after consumption of KCA1.

**Conclusion:** Our findings suggest that *Lactobacillus pentosus* KCA1 taken orally, lowers pro-inflammatory cytokine, IL-1 beta and decreases the relative abundance of BV-associated bacteria.

## INTRODUCTION

Bacterial vaginosis (BV) is under-reported, misdiagnosed and inappropriately treated in Nigeria^.[**1**]^ Our recent paper reported that a large proportion of women (45.1%) assumed that vaginal discharge is the most common symptom associated with BV and 41.7% stated that BV recurrence is a common denominator in their lives. On the Social burden of BV, 53.7% of women indicated that BV makes them avoid having sex while 29.5% avoid closeness with people, yet Nigerian clinicians still rely on antibiotics.^[**2**]^ Antibiotics reduces in considerable proportion both the healthy bacteria especially Lactobacilli and those associated with BV, thus the vaginal microecology and gut is adversely affected.^**[3]**^ It is widely acknowledged that Lactobacilli and other genera that produces some metabolites such as lactic acid, contributes to vaginal health by maintaining the low pH of the vagina.^**[4]**^ Although the etiology of BV is still unresolved, a recent metagenomics study has provided an insight into the phylogenetic diversity and species richness of the vagina and gut microbiome of reproductive age women with BV in some selected Nigerian women.^**[5]**^ The polymicrobial nature of BV is also believed to induce a pro-inflammatory environment, consisting of cytokines and toll-like receptor (TLR) ligands.^**[6]**^ In Nigeria, clinicians treat BV with antibiotics combinations especially Metronidazole or Clindamycin. However, the recurrence rate remains high. Failure of the antibiotics and recurrences common in BV are attributed to various reasons such as drug resistance, failure of the agents to penetrate and eradicate biofilms formed by BV associated organisms.^**[7]**^ The concept of replenishing the vaginal microbiota can be traced back to antiquity but it was reinvigorated four decades ago by delivering lactobacilli orally to repopulate the vagina.^[**8**]^ A potential probiotic organism, *Lactobacillus pentosus strains* KCA1 was isolated for the first time from the vagina of a healthy Nigerian women. Some microbiologists have scaled up their decades of research on few vaginal Lactobacillus strains as probiotics, thus, *Lactobacillus pentosus* KCAI was shown to produce biosurfactants, hydrogen peroxide (H_2_O_2_) and inhibit the intestinal and urogenital pathogens, ^**[9]**^ as well as exhibit varying degrees of acid bile tolerant.^**[10]**^

In order to establish the genetic capability and functional potentials, the full genome sequence of *Lactobacillus pentosus* KCA1 was determined in our previous study.^**[11]**^ *Lactobacillus pentosus* KCA1 was found to dedicate 121 genes to metabolism of cofactors and vitamins including five genes for biotin biosynthesis. The *L. pentosus* KCA1 genome also encodes putative phage defense systems including CRISPRs and abortive infection, novel toxin-antitoxin systems, and biosynthesis of an antibacterial peptide, a class V cyclic bacteriocin precursor, designated as pentocin KCA1 (penA). There are five complete two-component systems in KCA1 which includes histidine kinase hpk1 (KCA1_0030) and response regulator rrp1 (KCA1_0029) and two component response regulator TrxR (KCA1_2843), a transcriptional regulator of the AraC family. There have been multiple studies showing significant improvements in treating vaginal infections with probiotics versus traditional antibiotics treatments but so far only few strains have been clinically proven to be effective in particular to prevent BV and no study has been done to determine the impact of a single Lactobacillus strain on the vaginal and gut microbiome. It is important to note that Lactobacillus strains are specific in their action according to Shanahan ^**[12]**^ and Van *et al*.,^**[13]**^ denoting that one should not utilize data from one strain to infer to another untested strain that will provide the same benefit. It is critically important that *Lactobacillus pentosus KCA1*, which has been characterized at molecular level be tested clinically for probiotic use.

The genome provides a basis for its application to maintain a balanced urogenital health especially for women diagnosed with BV. The objectives of this study are three folds, first to determine the effects before and after 14 days oral feeding of *Lactobacillus pentosus* KCA1 on the vaginal and gut microbiota compositions of women diagnosed with BV. Second to measure the level of two proinflammatory cytokines IL-1 beta and IL-6 before and after KCA1 consumption and third to determine in silico, the relative abundance of microbial genes involved in metabolic functions using the 16S rRNA dataset.

## METHODS AND MATERIAL

### Ethical approval

Ethical approval was obtained from the research ethics review board of the faculty of health science and technology, Nnamdi Azikiwe University (NAUTH/CS/66/10/75/2017/046) and General hospital, Onitsha, in accordance with the Helsinki Declaration of 1975, as revised in 2000 (available at http://www.wma.net/e/policy/17-c_e.html). The protocol has been registered online at ClinicalTrials.gov with a unique identifier: NCT04329338.

### Study location and population

This study was conducted at General Hospital Onitsha, Anambra State. This hospital is the only government hospital at Onitsha and serves the neighbouring communities. One hundred and twenty women of reproductive age group who visited clinic at GHO were recruited for this study. The subjects were randomly selected after informed consent.

### Inclusion criteria are subjects with

age range 18 to 49 years, not on antibiotic therapy within the last one month and not menstruating at the time of inclusion. Subjects were **excluded if found to be** Pregnant, use of antibiotic medication, declined informed consent, menstruating and those that used douches, sprays, spermicides in the last 48 hours.

### Collection of Specimen

Two high vaginal swab were collected by a qualified Gynecologist with a non-lubricated sterile disposable plastic speculum and samples were collected from the posterior vaginal fornix using a Dacron swab stick.

### Microscopy and Nugent Score

Bacterial vaginosis was determined using Gram staining technique and Nugent criteria. One set of the high vaginal swab was used to prepare a dry vaginal smear which was stained by the Gram staining technique. The slides were examined using the 100x (oil immersion objective) and scored for BV using Nugent criteria. The slides were observed for presence of four different bacterial morphotypes: large Gram-positive rods (*Lactobacillus* spp. morphotypes); small Gram-variable rods (*G. vaginalis* morphotypes); small Gram-negative rods (*Bacteroides* spp. morphotypes); and curved Gram-variable rods (*Mobiluncus* spp. morphotypes). For *Lactobacillus*, scores range from 0-4; where 0 indicates that 30 or more organisms are identified and 4 indicate that no organisms are identified in the sample. For *Gardnerella* however, a score of 0 indicates that no organisms are identified and the highest score of 4 indicates that 30 or more organisms are identified. For *Mobiluncus*, scores range from 0-2 with a score of 2 indicating that 5 or more organisms are identified in the sample. At the end of scoring process, total scores ranging from 0 to 10 was computed. A total score of 7 or more was termed a case of bacterial vaginosis; a score of 4 to 6 was considered intermediate while a score of 0 to 3 was considered normal as standardized by Nugent.^**[14]**^

### Oral Administration of *Lactobacillus pentosus* KCA1

Seven women diagnosed with BV by Nugent score (7-10) were recruited to provide vaginal and gut sample before and after 14 days oral intake of 3 grams (2.5X108 cfu/g) of *Lactobacillus pentosus* KCA1 (Winclove probiotics, Amsterdam-The Netherlands) suspended in lukewarm water or dairy and taken daily for 14 days. Blood samples were also collected before and after 14 days of oral consumption of *Lactobacillus pentosus* KCA1.

### Extraction of bacterial DNA from vaginal swabs/stool samples and Sequencing of the amplified 16S rRNA region

Bacterial DNA was extracted from the vaginal swabs using an in-house protocol developed by Ubiome Inc. Briefly, samples were lysed using bead-beating, and DNA was extracted in a class 1000 clean room by a guanidine thiocyanate silica column-based purification method using a liquid-handling robot. PCR amplification of the 16S rRNA genes was performed with primers containing universal primers amplifying the V4 region (515F: GTGCCAGCMGCCGCGGTAA and 806R: GGACTACHVGGGTWTCTAAT). In addition, the primers contained Illumina tags and barcodes. Samples were barcoded with a unique combination of forward and reverse indexes allowing for simultaneous processing of multiple samples. PCR products were pooled, column-purified, and size-selected through microfluidic DNA fractionation. Consolidated libraries were quantified by quantitative real-time PCR using the Kapa Bio-Rad iCycler qPCR kit on a BioRad MyiQ before loading into the sequencer. Sequencing was performed in a pair-end modality on the Illumina NextSeq 500 platform rendering 2 ⨯150 bp pair-end sequences.

### Metagenomics sequence analysis

Raw sequence reads were demultiplexed using Illumina’s BCL2FASTQ algorithm. Reads were filtered using an average Q-score > 30. The paired-end sequence FASTQ reads were imported into MG-RAST pipeline for quality check (QC). EzBiocloud Microbiome Taxonomic Profile (MTP) pipeline,^**[15]**^ was employed for alpha and beta diversity estimation using PKSSU4.0 version database and Open reference UCLUST_MC2 for OTUs picking at 97% cut-off. Sequences were pre-screened using QIIME-UCLUST algorithms for at least 97% identity to ribosomal sequences from the RNA databases.^**[16]**^ Rarefication to 1000 reads per sample was employed to calculate microbial diversity. Alpha-diversity was calculated for species richness by ACE, Chao1 and Jackknife method, while diversity indexes were calculated by Shannon, Non-parametric Shannon and Simpson index. Principle coordinate analysis (PCoA) with Jensen-Shannon divergence distance metrices were used to evaluate beta diversity between vaginal and gut samples.^**[17]**^ Linear discriminant analysis (LDA) effect size (LEfSe) ^**[18]**^ was used to identify biologically and statistically significant differences in the OTU relative abundance. Phylogenetics Investigation of Communities by Reconstruction of Unobserved States (PICRUSt) was used to predict the metabolic function of the metagenomes from the 16S rRNA gene dataset,^**[19]**^ with reference to Kyoto Encyclopedia of Genes and Genomes (KEGG) Orthologs categorizations.^**[20]**^ Blood samples for cytokines were also collected before and after KCA1 consumption and processed with ELISA technique.

Statistical analysis: Student’s T test as obtained in Excel 2019 Office was used to test differences at p< 0.05 in the proportion of microbiota before and after oral intake of *L pentosus* KCA1.

## RESULTS

### Vaginal Microbiome composition before and after Consumption of *Lactobacillus pentosus* KCA1

The 16S rRNA data sets comprising data before and after consumption of *Lactobacillus pentosus* KCA1 were compared by using the Greengenes Version 13_8, (ftp://greengenes.microbio.me/greengenes_release/gg_13_5/gg_13_8_otus.tar.gz) in Illumina basespace algorithm as shown in **Figure 1**. At the phyla taxonomic categories, prior to consumption of *Lactobacillus pentosus* KCA1, twenty-two phyla were identified. Firmicutes (47.60%) were the most abundant, followed by Bacteriodetes (27.39%), Actinobacteria (17.10%), Fusobacteria (6.03%), Tenericutes (1.38%), Proteobacteria (0.44%), Acidobacteria (0.01%) and others as shown **Figure 2**. After 14 days consumption of *Lactobacillus pentosus* KCA1, twenty-two phyla were also identified with obliteration of two phyla (*Armatimonadetes* and *Planctomycetes*) and introduction of two new phyla (*Elusimicrobia* and *Nitrospirae*). Firmicutes (46.68%) were the most abundant, followed by Bacteriodetes (26.31%), Actinobacteria (18.01%), Fusobacteria (7.29%), Tenericutes (0.97%), Proteobacteria (0.66%), Acidobacteria (0.01%) and others as presented in **Figure 2**.

**Figure 1:**
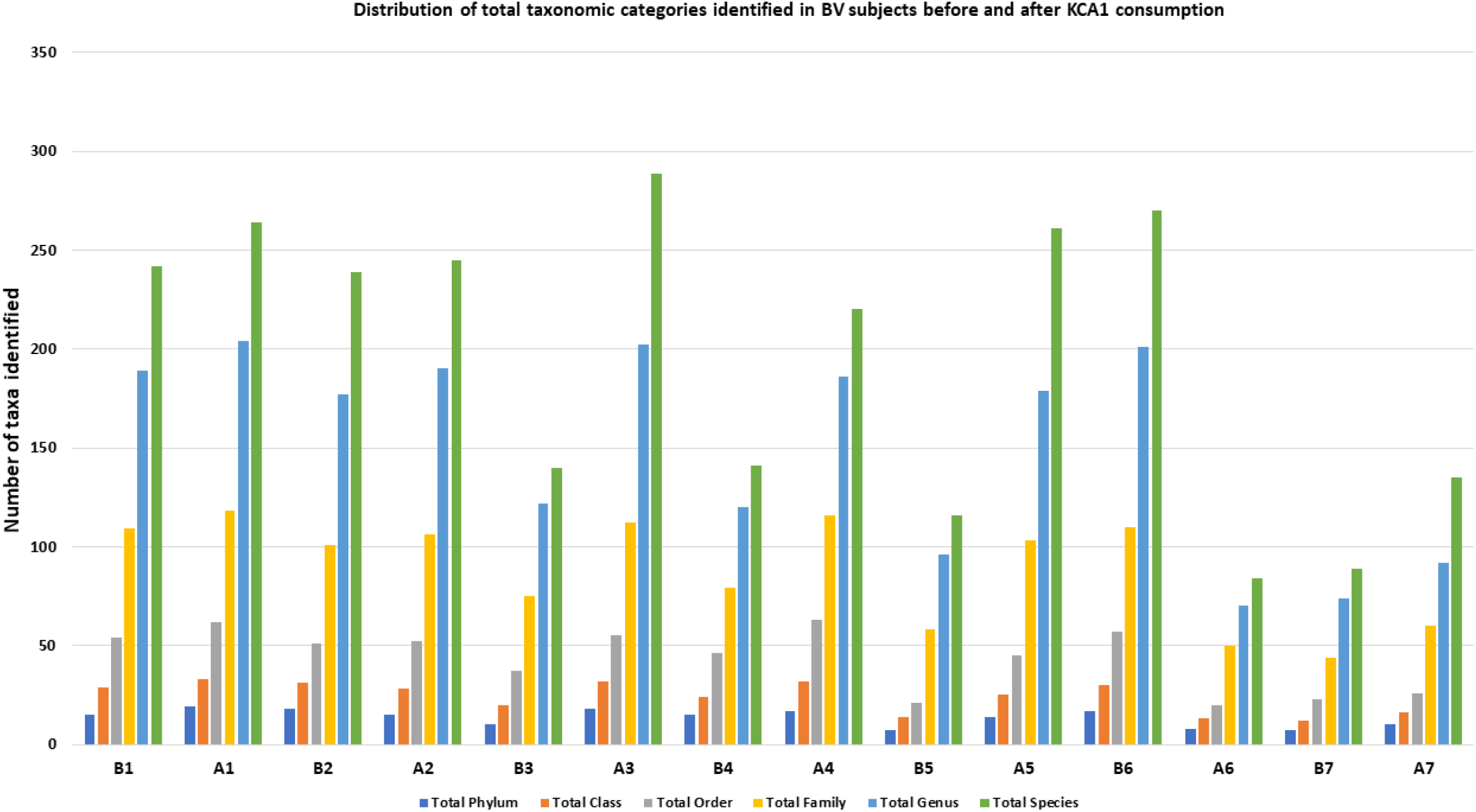
Distribution of total taxonomic categories in BV subjects before and after KCA1 consumption.

**Figure 2:**
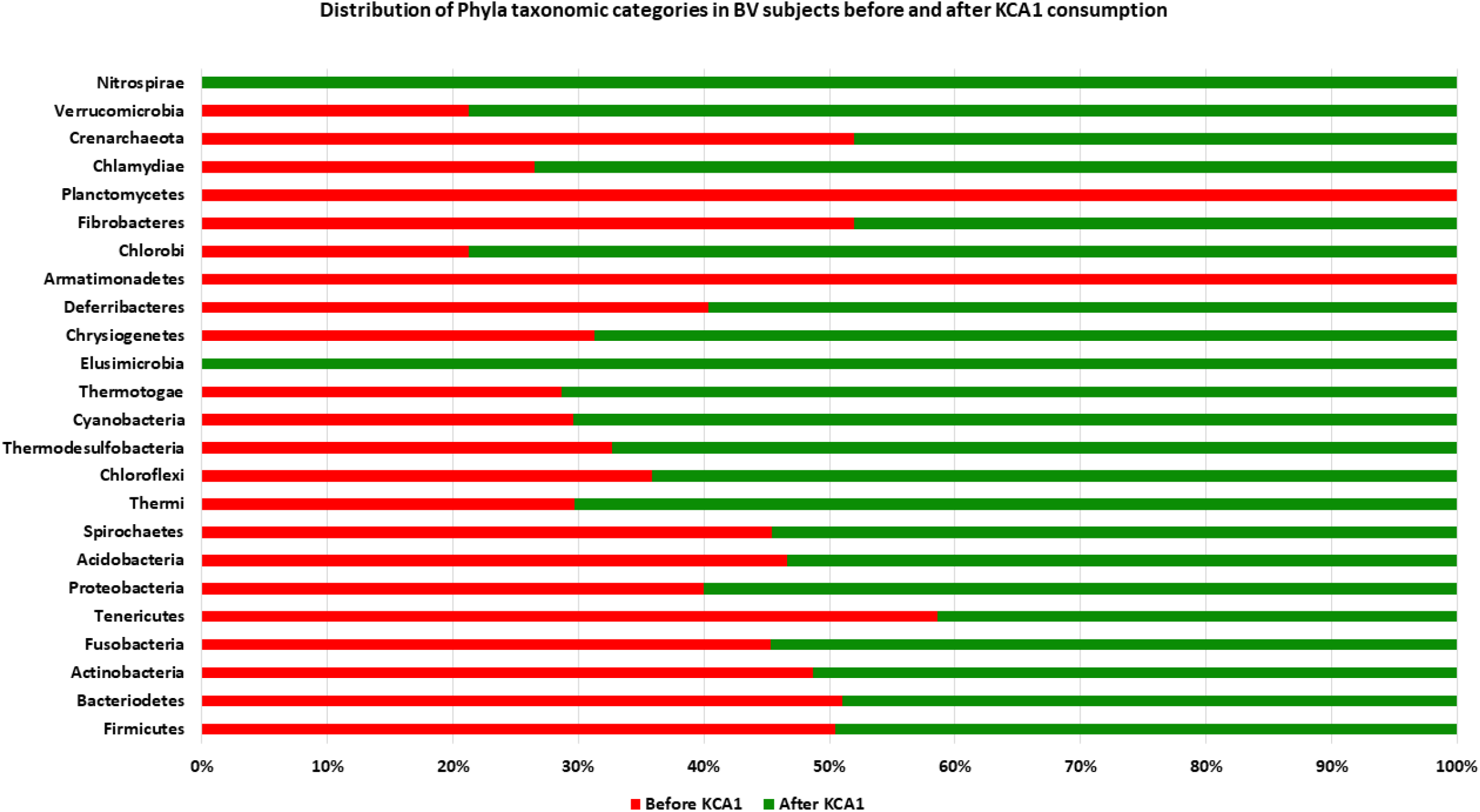
Distribution of Phyla taxonomic categories identified in BV subjects before and after *L pentosus* KCA1 consumption.

In addition, EzBioCloud Microbiome Taxonomic Profile (MTP) was used to determine alpha diversity including species richness and diversity index. On average, there was no significant difference (P=0.749) in the species richness-alpha indices typified by ACE, CHAO1, Jackknife, and number of OTUs. Diversity index such as NPShannon and Shannon (P=0.749) were not significant. Similarly, Simpson and Phylogenetic diversity (P=0.565) were not significant before and after KCA1 consumption (**Figure 3**). The beta diversity showed different clustering positions with Bray-Curtis Principal Coordinates Analysis (**Figure 4**). However, at individual taxonomic categories, there was a significant difference in the relative abundance of the phyla proteobacteria, actinobacteria and some genera associated with bacterial vaginosis observed before and after KCA1 feeding. For example, there was a 23.29% decrease in the genus Gardnerella, Porphyromonas (26.39%), Prevotella (16.1%), Gemella (52.07%), Veillonella (54.17%), Atopobium (43.91%), Ureaplasma (56.11%), and Peptostreptococcus decreased by 59.72%. with a corresponding 56.09% increase of Lactobacillus genera (**Figure 5**)

**Figure 3:**
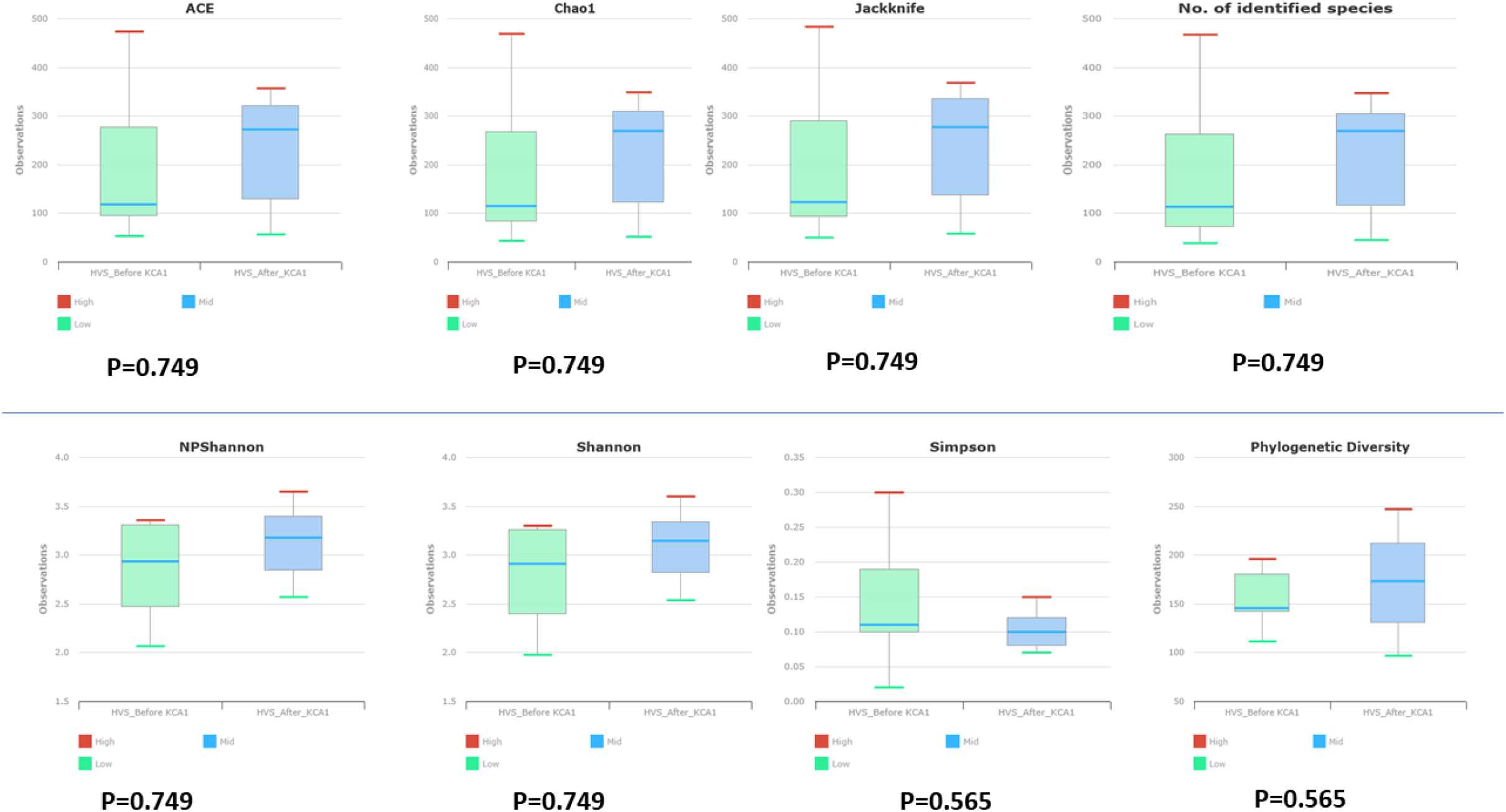
Alpha diversity index showing the species richness as determined by EzBioCloud MTP pipeline.

**Figure 4:**
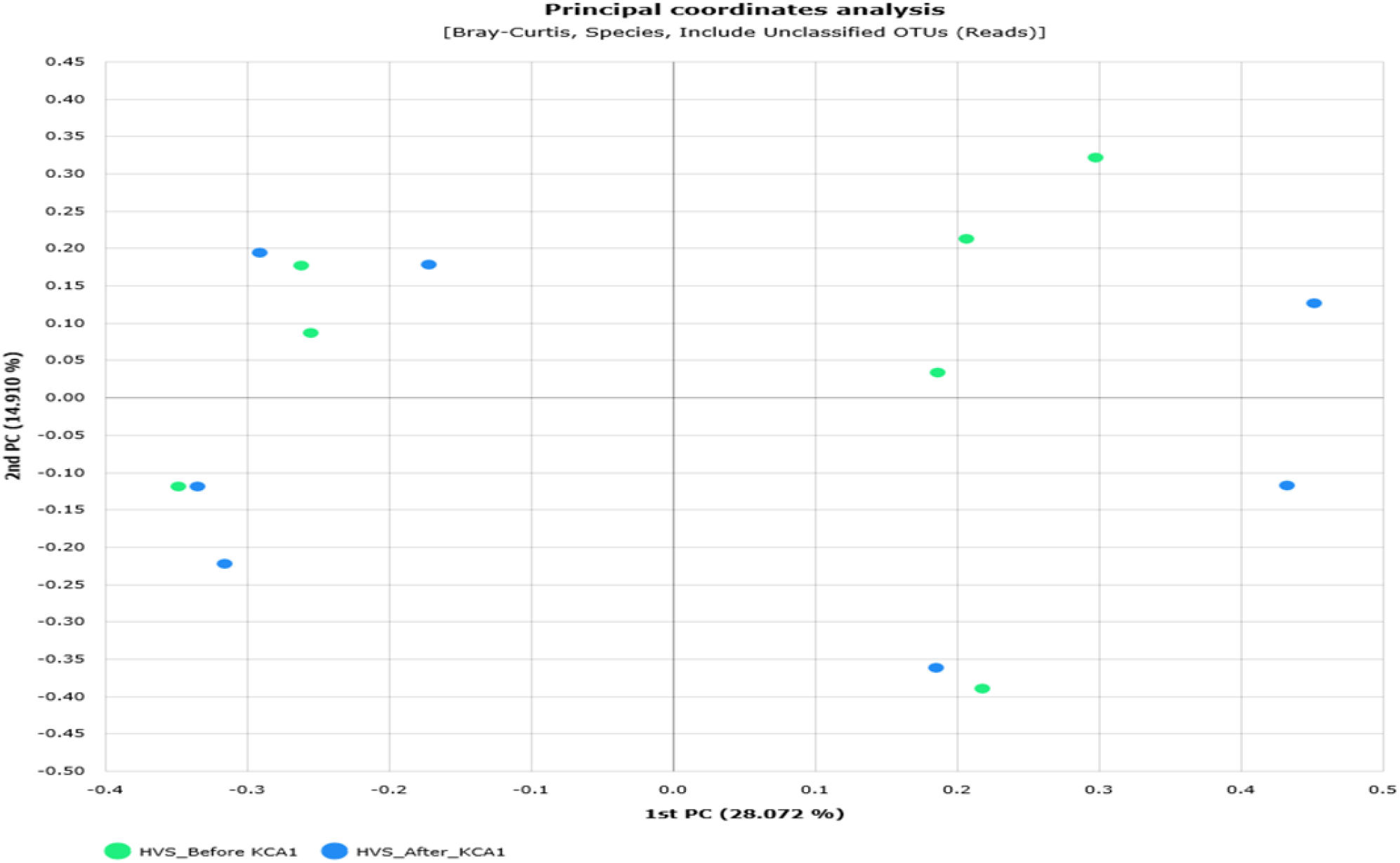
Beta diversity showing different clustering positions before and after *L pentosus* KCA1 with Bray-Curtis Principal Coordinates Analysis.

**Figure 5:**
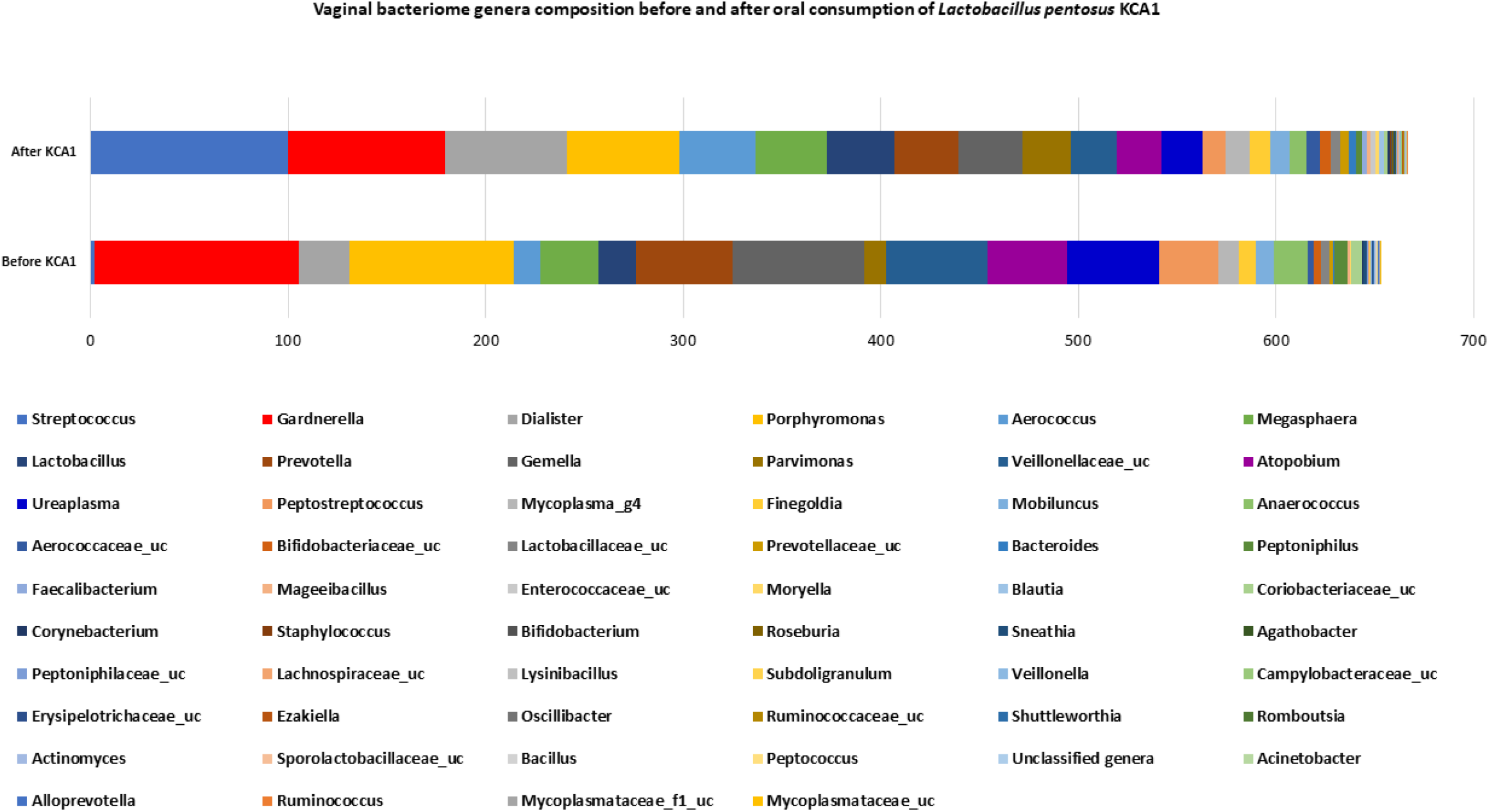
Vaginal bacteriome genera compositions before and after oral consumption of *Lactobacillus pentosus* KCA1.

At the species taxonomic level, some bacterial organisms associated with BV decreased with higher proportions as shown in **Figure 6**. Interestingly, 113 (37%) species were exclusively identified in the vagina of the subjects after KCA1 consumption. Notable species found after KCA1 were *Lactobacillus pentosus, Lactobacillus reuteri, Lactobacillus crispatus, Lactobacillus gasseri, Blautia wexlerae, Roseburia cecicola, Roseburia inulinivorans*, and *Faecalibacterium spp*.

**Figure 6:**
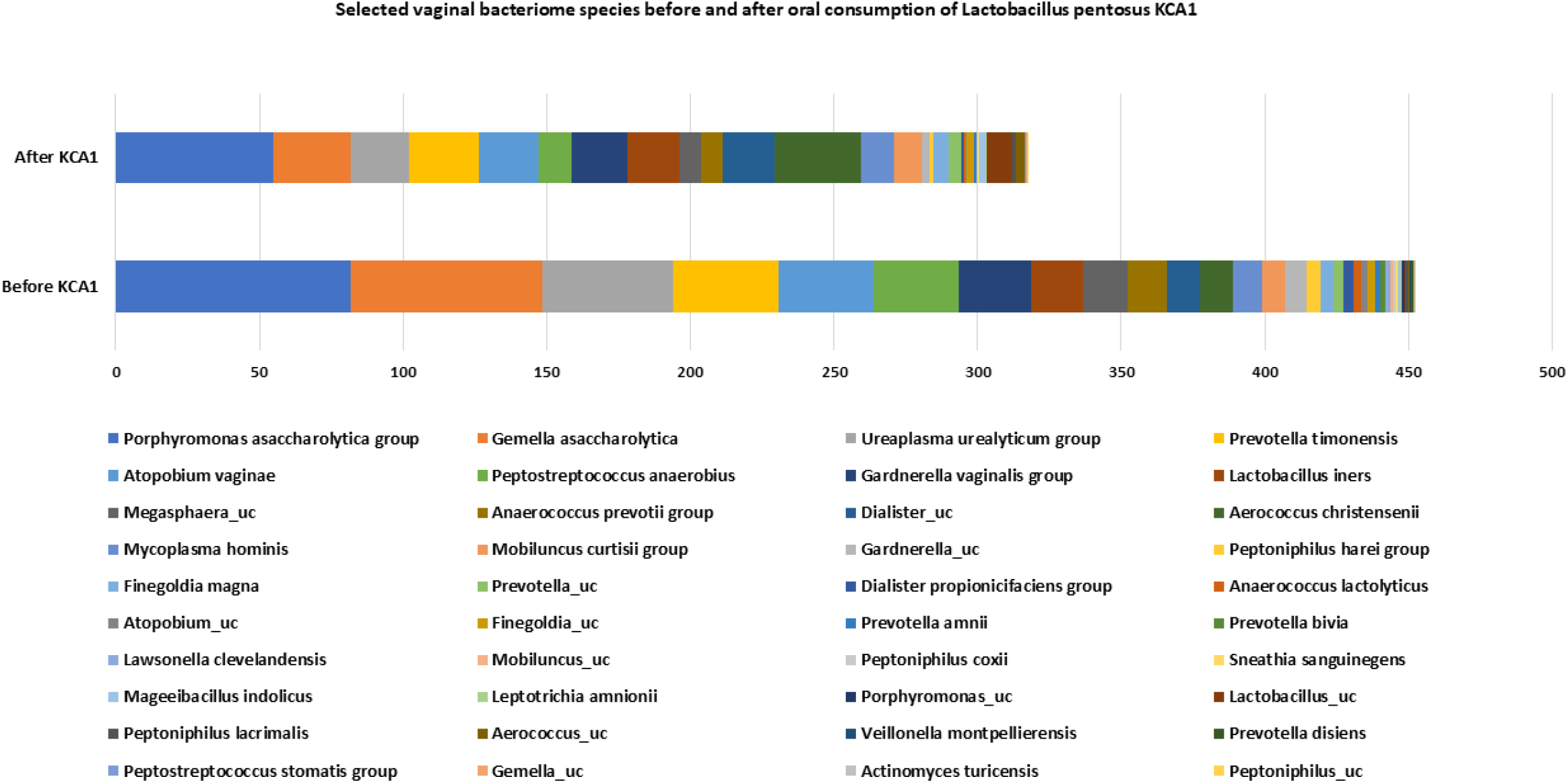
Selected vaginal bacteriome species before and after oral consumption of *Lactobacillus pentosus* KCA1.

### Gut microbiome composition before and after Consumption of *Lactobacillus pentosus* KCA1

The Gut microbiome before and after oral feeding of KCA1 rarefaction curve is presented in **Figure 7**. Microbiota composition at the genera taxonomic level shows that *Lactobacillus pentosus* KCA1 led to an increase in the proportion of some genera. For example, there was a 41.32% increase in *Faecalibacterium, Bacteroides* (29.94%), *Escherichia* (64.36%), *Clostridium* (3.93%), *Parasutterella* (3.97%), *Ruminococcus* (1.86%), *Streptococcus* (2.69%), *Lactobacillus* (3.20%), *Enterococcus* (0.22%), *Akkermansia* (0.17%) and *Butyricimonas* (0.74%). Conversely, there was a decrease in the relative abundance of some genera associated with bacterial vaginosis (**Figure 8**). *Alloprevotella* decreased by 18.05%, *Prevotella* (1.6%), *Haemophilus* (8.08%), *Intestinibacter* (2.63%), *Clostridium*_g24 (0.19%), *Coprococcus* (5.13%), *Prevotellaceae*_uc (0.64%), *Fusicatenibacter* (8.33%), *Megasphaera* (0.54%), *Porphyromonas* (0.16%), *Veillonella* (0.35%), *Actinomyces* (0.02%), *Peptostreptococcus* (0.12%), *Streptococcaceae*_uc (0.15%), *Acinetobacter* (2.8%), *Anaerococcus* (0.04%), *Aeromonas* (0.17%), *Eggerthella* (0.05%), *Neisseria* (0.05%), *Campylobacter* (0.02%), *Staphylococcus* (0.02%), *Clostridium*_g21 (0.08%), *Atopobium* (0.02%) and *Gardnerella* (0.09%). Some genera that were identified before oral intake of KCA1 appears to be absent after KCA1 consumption. The Venn diagram plot shows that 67.2% of the genera were commonly present before and after, while 18.7% were exclusively found after KCA1 oral intake and 14.2% were exclusively found pre-KCA1 consumption (**Figure 9**). Such genera include but not limited to *Achromobacter, Actinobacillus*_g1, *Actinobaculum, Aggregatibacter, Alcaligenes, Alishewanella, Anaerofustis, Anaerotaenia, Arcobacter, Burkholderia, Mobiluncus, Pediococcus, Pelomonas, Peptococcus, Slackia, Sphingobacterium, Ureaplasma, Vallotia* and *Vibrio*. Some genera that occurred below 1% relative abundance are presented in **Figure 10**.

**Figure 7:**
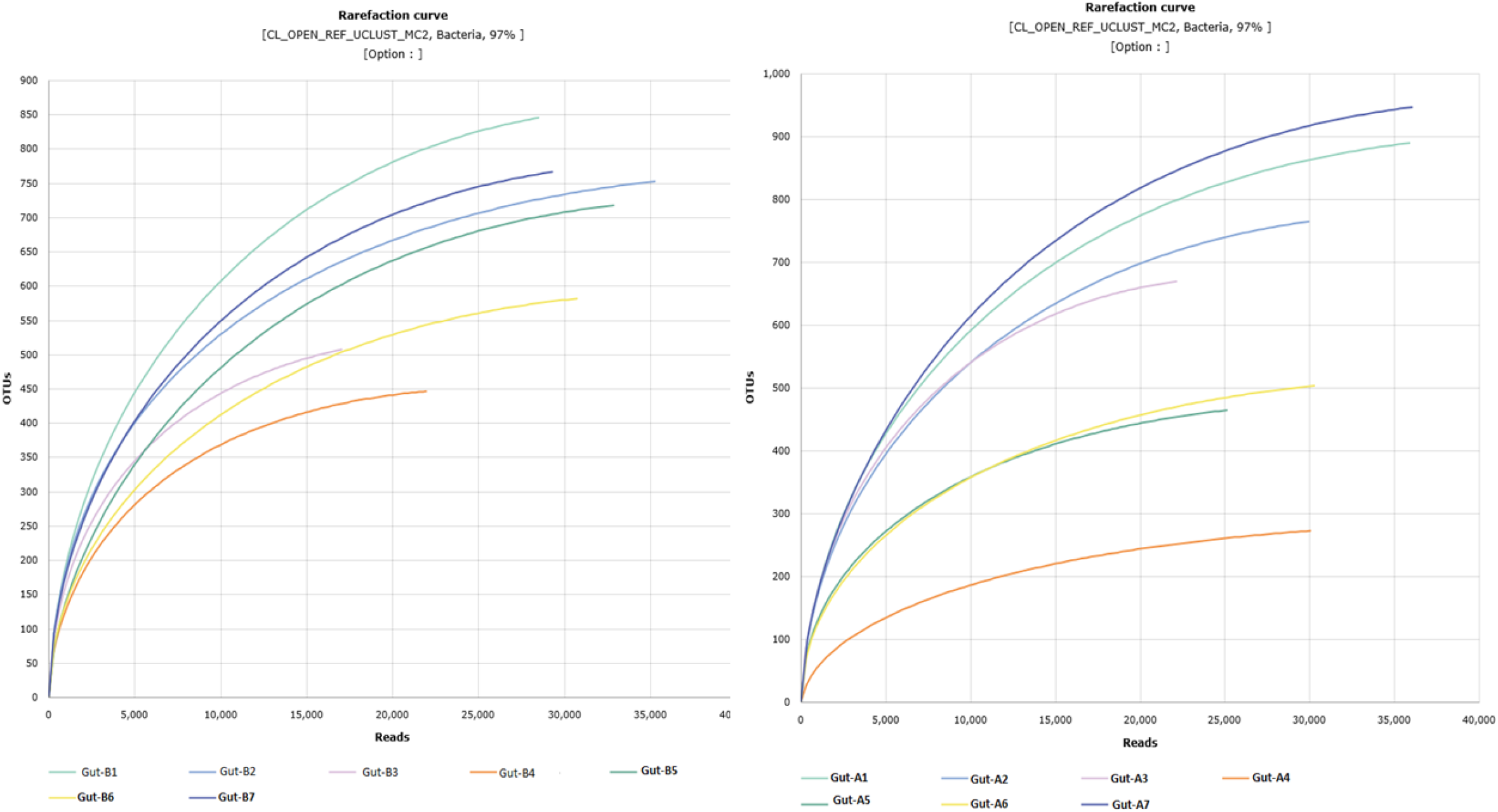
Rarefaction curve showing the number of reads from gut samples associated with the number of operational taxonomic units (OTUs)

**Figure 8:**
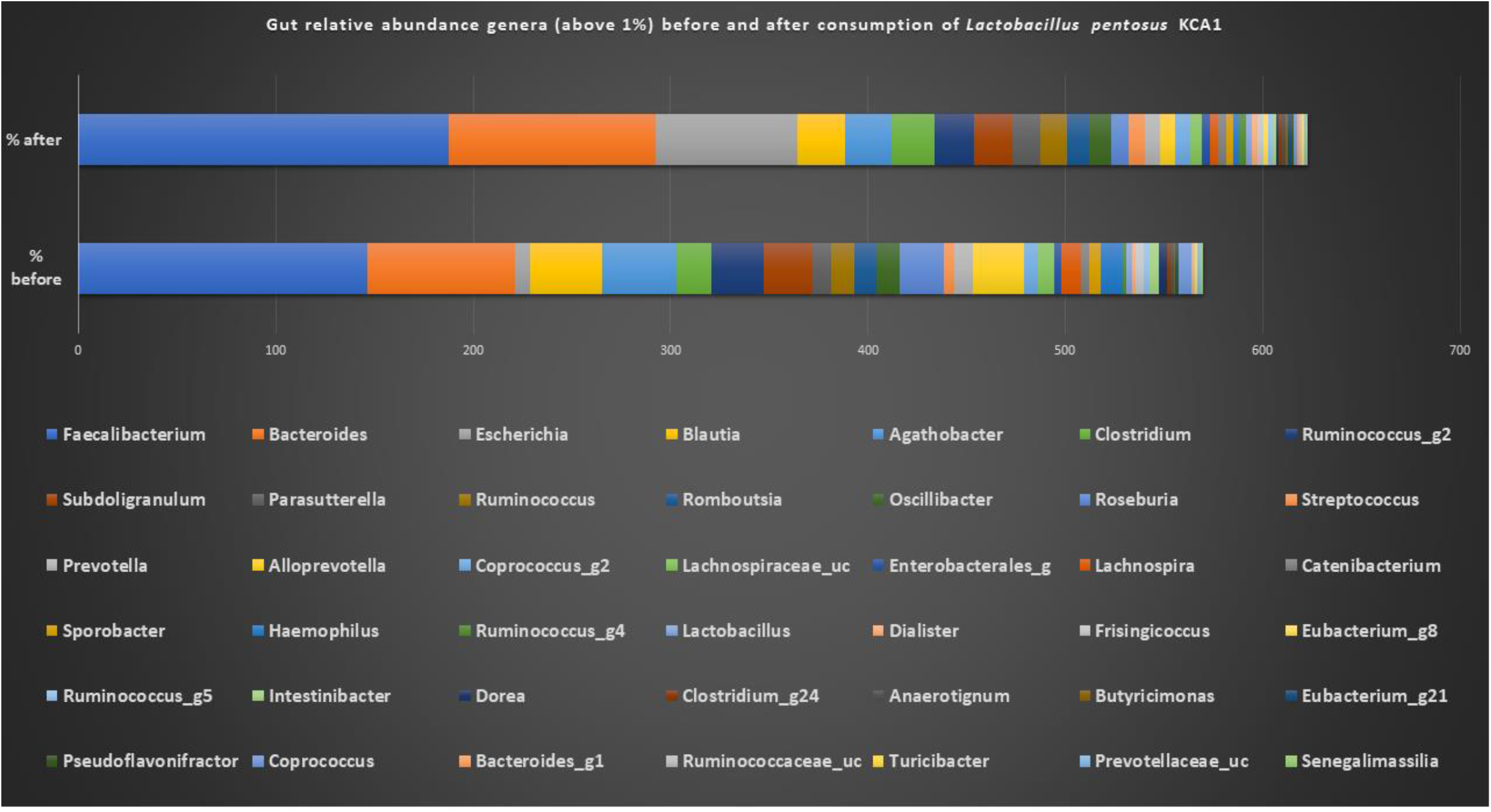
Gut relative abundance genera (above 1%) before and after consumption of *Lactobacillus pentosus* KCA1.

**Figure 9:**
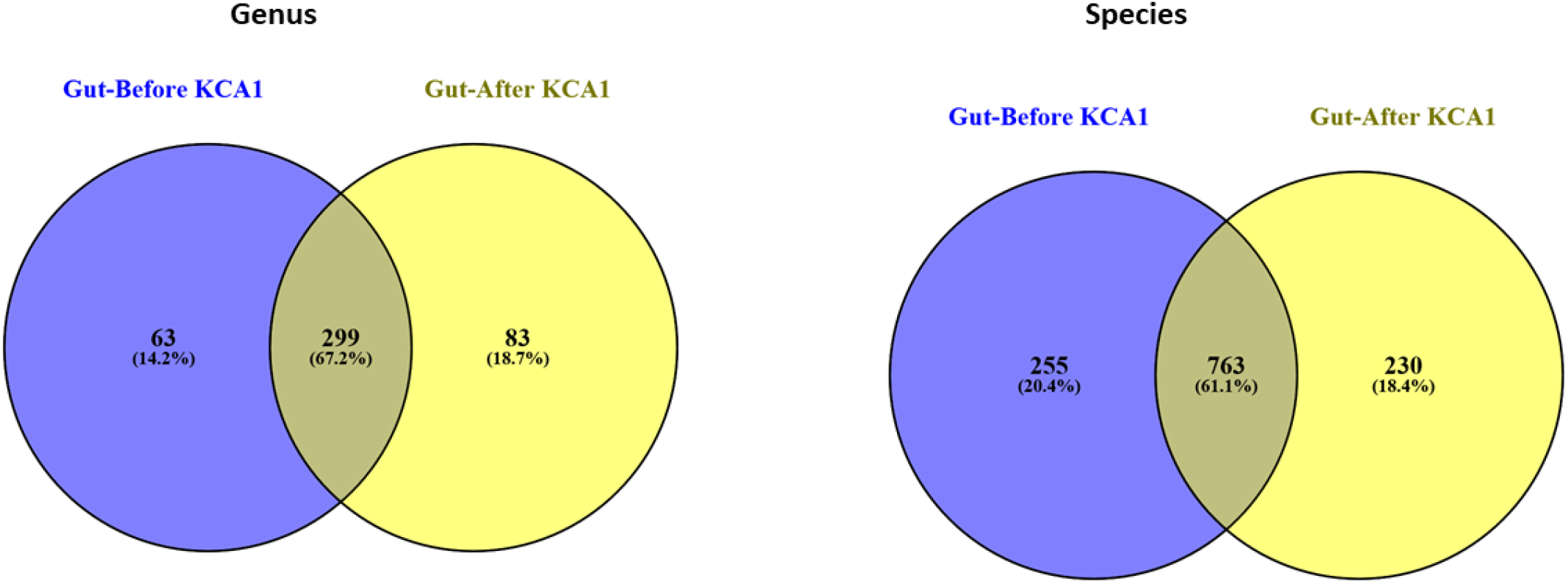
Venn diagram showing the number (%) of genus and species identified in the gut that were common and exclusive before and after oral intake of *L. pentosus* KCA1.

**Figure 10:**
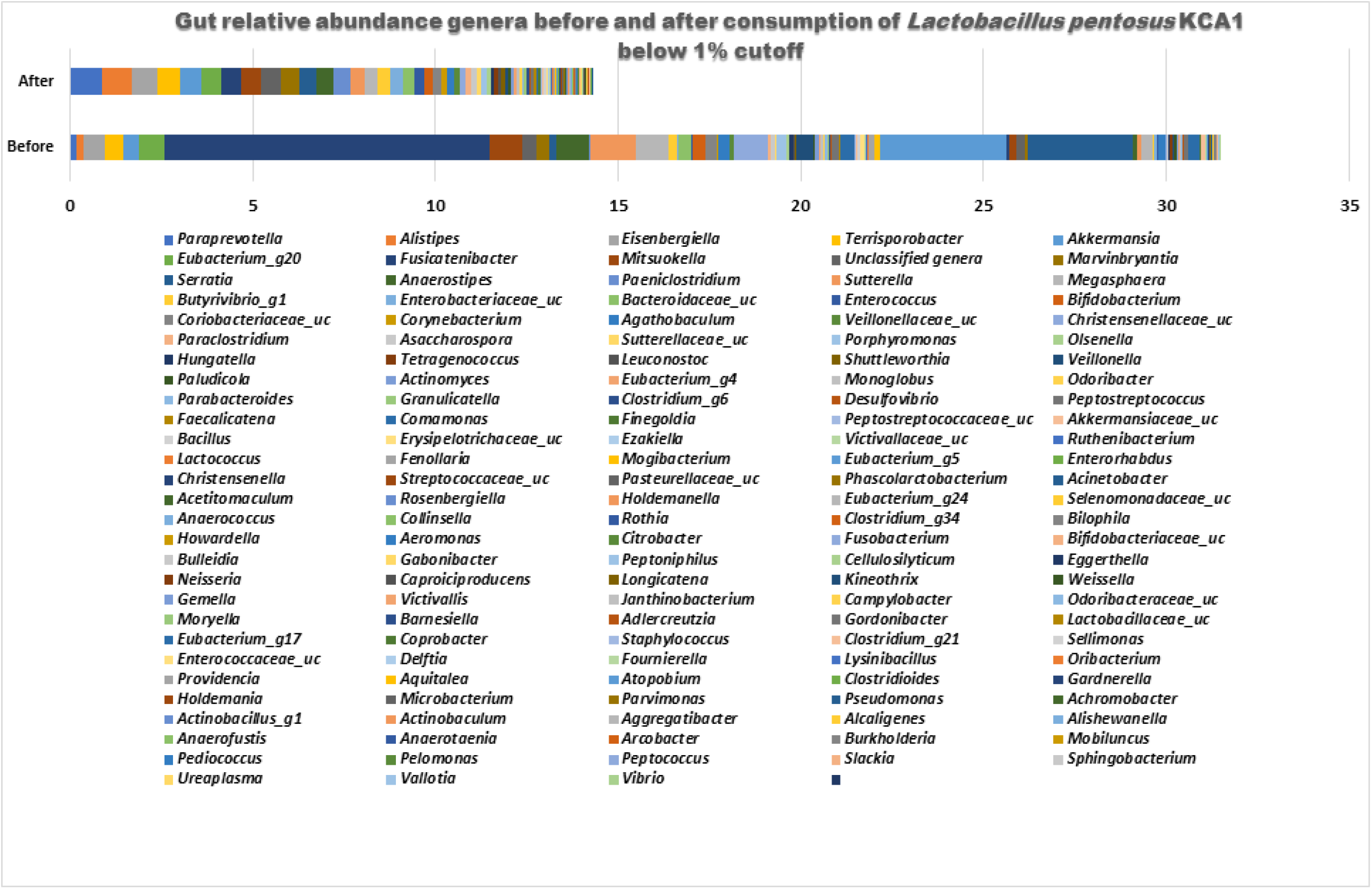
Gut relative abundance genera (below 1% cutoff) before and after consumption of *Lactobacillus pentosus* KCA1.

### Metabolic Functional Prediction using the 16S rRNA Marker Genes

The results from PICRUSt indicated that certain bacterial metabolic functional genes were upregulated in the vagina after consumption of *Lactobacillus pentosus* KCA1. For example, metabolic pathways for amino acid, alanine was significantly upregulated (p= 0.008809) with LDA effect size of 3.445098. Similarly, polar amino acid transport system substrate-binding protein with gene ortholog -K02030, was significantly increased (LDA effect size= 2.364761; and *P*= 0.035006) (**Figure 11)**. There was significant difference in the predicted relative abundance of microbial gene pathways related to degradation of aromatic compounds (ko01220) with LDA effect size = 2.571179 and P= 0.047645. There was also high abundance of two-component system sensor histidine kinase; two-component system, OmpR family, lantibiotic biosynthesis sensor histidine kinase NisK/SpaK and redox-sensing transcriptional regulator. There were acquisitions of bacterial metabolic genes related to defence systems such as CRISPR-associated protein Cmr1, and DNA mismatch repair protein MSH3, after consumption of KCA1 in the vagina (**Table 1**)

**Table 1:** Bacterial metabolic functional genes predicted with PICRUSt using EzBiocloud pipeline.

| Gene Ortholog | Definition | p-value | p-value (FDR) | Vagina Before KCA1 | Vagina After KCA1 |
| --- | --- | --- | --- | --- | --- |
| K03367 | D-alanine--poly(phosphoribitol) ligase subunit 1 | 0.002676 | 0.002675786 | 0.011612774 | 0.023490698 |
| K10254 | oleate hydratase | 0.002676 | 0.002676023 | 0.005788293 | 0.019083057 |
| K02160 | acetyl-CoA carboxylase biotin carboxyl carrier protein | 0.006011 | 0.006012273 | 0.008628376 | 0.021727122 |
| K07469 | aldehyde oxidoreductase | 0.007797 | 0.007799211 | 1.69334E-06 | 9.58652E-05 |
| K14007 | protein transport protein SEC24 | 0.007797 | 0.007799901 | 1.69334E-06 | 4.38724E-05 |
| K08899 | **lemur tyrosine kinase 3** | 0.009162 | 0.009166487 | 0 | 8.28136E-05 |
| K05906 | **prenylcysteine oxidase / farnesylcysteine lyase** | 0.009162 | 0.009167297 | 0 | 3.67334E-05 |
| K18357 | **phenylglyoxylate dehydrogenase gamma subunit** | 0.009162 | 0.009168108 | 0 | 6.20031E-05 |
| K10694 | **E3 ubiquitin-protein ligase ZNRF1/2** | 0.009162 | 0.009168919 | 0 | 8.80003E-05 |
| K22017 | **mucin-4** | 0.009162 | 0.009169729 | 0 | 0.000108002 |
| K08353 | **thiosulfate reductase electron transport protein** | 0.009162 | 0.00917054 | 0 | 7.4644E-05 |
| K13258 | **2-hydroxyisoflavanone dehydratase** | 0.009162 | 0.009171351 | 0 | 3.49778E-05 |
| K16093 | **bacitracin synthase 1** | 0.009162 | 0.009172163 | 0 | 0.000138736 |
| K02763 | PTS system, D-glucosamine-specific IIA component | 0.011593 | 0.011607835 | 1.15334E-05 | 0.000206032 |
| K10049 | CCAAT/enhancer binding protein (C/EBP), gamma | 0.011593 | 0.011608862 | 2.88335E-06 | 9.26212E-05 |
| K18841 | mRNA interferase ChpB | 0.011593 | 0.011609889 | 2.88335E-06 | 0.000106026 |
| K18890 | ATP-binding cassette, subfamily B, multidrug efflux pump | 0.012716 | 0.012735379 | 0.011796924 | 0.028984128 |
| K07482 | transposase, IS30 family | 0.012716 | 0.012736506 | 0.07559319 | 0.168850196 |
| K01961 | acetyl-CoA carboxylase, biotin carboxylase subunit | 0.012716 | 0.012737634 | 0.012246525 | 0.023640415 |
| K06501 | CD68 antigen | 0.016672 | 0.016701182 | 1.11901E-05 | 0.000221225 |
| K07260 | **zinc D-Ala-D-Ala carboxypeptidase** | 0.018086 | 0.018120082 | 0.008296247 | 0.03152758 |
| K07795 | putative tricarboxylic transport membrane protein | 0.018086 | 0.018123291 | 0.007612762 | 0.002018814 |
| K02236 | leader peptidase (prepilin peptidase) / N-methyltransferase | 0.018086 | 0.018124895 | 0.00592522 | 0.020946159 |
| K18592 | gamma-glutamyltranspeptidase / glutathione hydrolase / leukotriene-C4 hydrolase | 0.019554 | 0.019597171 | 1.69334E-06 | 1.812E-05 |
| K10674 | ectoine hydroxylase | 0.019554 | 0.019598907 | 7.14861E-05 | 7.24365E-07 |
| K14325 | RNA-binding protein with serine-rich domain 1 | 0.023756 | 0.023812798 | 2.52825E-05 | 0.000102635 |
| K16267 | zinc and cadmium transporter | 0.023756 | 0.023814907 | 0.00034831 | 2.82608E-05 |
| K15659 | **arthrofactin-type cyclic lipopeptide synthetase B** | 0.025046 | 0.025110532 | 0 | 3.98252E-05 |
| K14832 | **ribosome biogenesis protein MAK21** | 0.025046 | 0.025112757 | 0 | 4.61488E-05 |
| K21368 | **UDP-Gal:alpha-L-Fuc-1,2-beta-Gal-1,3-alpha-GalNAc-1,3-alpha-GalNAc-diphosphoundecaprenol alpha-1,3-galactosyltransferase** | 0.025046 | 0.025114981 | 0 | 4.33456E-05 |
| K20790 | **nucleoside diphosphate kinase homolog 5** | 0.025046 | 0.025117207 | 0 | 3.16051E-05 |
| K13703 | abhydrolase domain-containing protein 11 | 0.025046 | 0.025119433 | 0.000258153 | 0 |
| K17639 | ral guanine nucleotide dissociation stimulator-like 3 | 0.025046 | 0.025121659 | 0.000527126 | 0 |
| K12526 | **bifunctional diaminopimelate decarboxylase / aspartate kinase** | 0.025046 | 0.025123885 | 0 | 3.16051E-05 |
| K04334 | **major curlin subunit** | 0.025046 | 0.025126112 | 0 | 1.98246E-05 |
| K14486 | **auxin response factor** | 0.025046 | 0.025128339 | 0 | 4.09081E-05 |
| K06487 | **integrin alpha V** | 0.025046 | 0.025130567 | 0 | 4.22859E-05 |
| K09482 | **glutamyl-tRNA(Gln) amidotransferase subunit D** | 0.025046 | 0.025132795 | 0 | 5.87962E-05 |
| K20811 | **inulosucrase** | 0.025046 | 0.025135024 | 0 | 0.000242737 |
| K16709 | **amylovoran biosynthesis protein AmsF** | 0.025046 | 0.025137252 | 0 | 1.66628E-05 |
| K11080 | **mannopine transport system ATP-binding protein** | 0.025046 | 0.025139482 | 0 | 5.54126E-05 |
| K07357 | **type 1 fimbriae regulatory protein FimB** | 0.025046 | 0.025141711 | 0 | 1.98246E-05 |
| K10620 | **2,3-dihydroxy-2,3-dihydro-p-cumate dehydrogenase** | 0.025046 | 0.025143941 | 0 | 0.000105834 |
| K20370 | **phosphoenolpyruvate carboxykinase (diphosphate)** | 0.025046 | 0.025146172 | 0 | 3.03629E-05 |
| K10022 | arginine/ornithine transport system substrate-binding protein | 0.025046 | 0.025148403 | 5.19604E-05 | 0 |
| K07314 | **serine/threonine protein phosphatase 2** | 0.025046 | 0.025150634 | 0 | 1.85825E-05 |
| K12397 | **AP-3 complex subunit beta** | 0.025046 | 0.025152866 | 0 | 0.000111236 |
| K21886 | **ArsR family transcriptional regulator, nickel/cobalt-responsive transcriptional repressor** | 0.025046 | 0.025155098 | 0 | 3.16051E-05 |
| K01093 | **4-phytase / acid phosphatase** | 0.025046 | 0.02515733 | 0 | 1.98246E-05 |
| K07061 | **CRISPR-associated protein Cmr1** | 0.025046 | 0.025159563 | 0 | 1.09144E-05 |
| K16359 | **netrin-G2** | 0.025046 | 0.025161796 | 0 | 0.000147139 |
| K09588 | **cytochrome P450 family 90 subfamily A polypeptide 1** | 0.025046 | 0.02516403 | 0 | 0.000175294 |
| K13415 | **protein brassinosteroid insensitive 1** | 0.025046 | 0.025166264 | 0 | 0.000123999 |
| K08736 | **DNA mismatch repair protein MSH3** | 0.025046 | 0.025168498 | 0 | 3.98252E-05 |
| K10666 | **E3 ubiquitin-protein ligase RNF5** | 0.025046 | 0.025170733 | 0 | 0.000143705 |
| K14837 | **nucleolar protein 12** | 0.025046 | 0.025172968 | 0 | 5.63805E-05 |
| K17890 | **autophagy-related protein 16-1** | 0.025046 | 0.025175204 | 0 | 7.05349E-05 |
| K18222 | **arylsulfatase E** | 0.025046 | 0.02517744 | 0 | 4.74866E-05 |
| K11693 | peptidoglycan pentaglycine glycine transferase (the first glycine) | 0.025347 | 0.025482408 | 0.003741601 | 0.008570039 |
| K01926 | redox-sensing transcriptional repressor | 0.025347 | 0.025484672 | 0.016331308 | 0.030946666 |
| K03740 | **D-alanine transfer protein** | 0.025347 | 0.025486936 | 0.013102146 | 0.027267324 |
| K11616 | malate:Na<sup>+</sup> symporter | 0.025347 | 0.025489201 | 0.003303801 | 0.011965665 |
| K07467 | **phage replication initiation protein** | 0.025347 | 0.025491466 | 0.004323412 | 0.016619946 |
| K03355 | anaphase-promoting complex subunit 8 | 0.025347 | 0.025493731 | 0.000753232 | 0.000311673 |
| K01126 | glycerophosphoryl diester phosphodiesterase | 0.025347 | 0.025495997 | 0.026346323 | 0.048183715 |
| K01728 | pectate lyase | 0.025347 | 0.025498263 | 0.004898644 | 0.013124259 |
| K07487 | **transposase** | 0.025347 | 0.025500529 | 0.018114649 | 0.258579872 |
| K10047 | inositol-phosphate phosphatase / L-galactose 1-phosphate phosphatase | 0.028286 | 0.028459509 | 5.7667E-06 | 0.000119604 |
| K21801 | indoleacetamide hydrolase | 0.028286 | 0.028462039 | 0.000305014 | 1.44873E-06 |
| K10624 | E3 ubiquitin-protein ligase RBBP6 | 0.033347 | 0.033557197 | 1.32366E-05 | 0.000178172 |
| K20098 | DNA excision repair protein ERCC-6-like 2 | 0.033347 | 0.033560181 | 7.43275E-06 | 0.000118784 |
| K03357 | anaphase-promoting complex subunit 10 | 0.033347 | 0.033563165 | 1.82381E-05 | 0.000217177 |
| K21672 | 2,4-diaminopentanoate dehydrogenase | 0.033347 | 0.03356615 | 2.3718E-05 | 0.000151075 |
| K02852 | UDP-N-acetyl-D-mannosaminouronate:lipid I N-acetyl-D-mannosaminouronosyltransferase | 0.034435 | 0.034664834 | 2.07059E-05 | 4.80906E-05 |
| K06019 | pyrophosphatase PpaX | 0.035006 | 0.035242333 | 0.001494945 | 0.009196995 |
| K03695 | ATP-dependent Clp protease ATP-binding subunit ClpB | 0.035006 | 0.035245468 | 0.100888013 | 0.079081389 |
| K00712 | poly(glycerol-phosphate) alpha-glucosyltransferase | 0.035006 | 0.035248604 | 0.007927823 | 0.020564455 |
| K22305 | phosphoserine phosphatase | 0.035006 | 0.03525174 | 0.0027264 | 0.013205239 |
| K02049 | NitT/TauT family transport system ATP-binding protein | 0.035006 | 0.035254877 | 0.089892498 | 0.112688192 |
| K00887 | undecaprenol kinase | 0.035006 | 0.035258015 | 0.009129932 | 0.022865891 |
| K01198 | xylan 1,4-beta-xylosidase | 0.035006 | 0.035261153 | 0.005169175 | 0.010305476 |
| K07483 | transposase | 0.035006 | 0.035264291 | 0.091623295 | 0.250486331 |
| K07171 | mRNA interferase MazF | 0.035006 | 0.03526743 | 0.010735722 | 0.024242624 |
| K01607 | 4-carboxymuconolactone decarboxylase | 0.035006 | 0.03527057 | 0.03435543 | 0.049065032 |
| K07717 | two-component system, sensor histidine kinase YcbA | 0.035006 | 0.03527371 | 0.001835765 | 0.010390348 |
| K17462 | putative AdoMet-dependent methyltransferase | 0.035006 | 0.035276851 | 0.003619675 | 0.007277721 |
| K02030 | polar amino acid transport system substrate-binding protein | 0.035006 | 0.035279992 | 0.1493949 | 0.195517253 |
| K16342 | cytosolic phospholipase A2 | 0.04019 | 0.040508252 | 2.04489E-05 | 0.000114601 |
| K15044 | Arf-GAP domain and FG repeats-containing protein 1 | 0.04019 | 0.04051186 | 2.88335E-06 | 3.81121E-05 |
| K19673 | tetratricopeptide repeat protein 21B | 0.04019 | 0.040515469 | 3.38667E-06 | 0.000106309 |
| K10820 | monosaccharide-transporting ATPase | 0.04019 | 0.040519078 | 4.51824E-06 | 8.73693E-05 |
| K22395 | cinnamyl-alcohol dehydrogenase | 0.04019 | 0.040522687 | 4.88522E-05 | 7.46067E-06 |
| K20324 | mutacin VI | 0.042812 | 0.043171024 | 1.85117E-05 | 9.70858E-05 |
| K08052 | neurofibromin 1 | 0.042812 | 0.04317487 | 0.000550575 | 1.57543E-05 |
| K21140 | [CysO sulfur-carrier protein]-S-L-cysteine hydrolase | 0.04667 | 0.04706947 | 2.88844E-05 | 0.000274593 |
| K05291 | phosphatidylinositol glycan, class S | 0.04667 | 0.047073665 | 2.5701E-05 | 0.0003826 |
| K08312 | ADP-ribose diphosphatase | 0.04667 | 0.04707786 | 5.14749E-05 | 0.000140524 |
| K13446 | disease resistance protein | 0.04667 | 0.047082057 | 2.5701E-05 | 0.000109775 |
| K00073 | ureidoglycolate dehydrogenase (NAD+) | 0.04667 | 0.047086254 | 0.000114001 | 0.000436769 |
| K15331 | tRNA (uracil-5-)-methyltransferase | 0.047401 | 0.047828057 | 5.10439E-05 | 0.000160251 |
| K00889 | 1-phosphatidylinositol-4-phosphate 5-kinase | 0.047645 | 0.04807872 | 0.001242012 | 0.009247603 |
| K07149 | uncharacterized protein | 0.047645 | 0.048083007 | 0.004262954 | 0.008282736 |
| K14731 | epsilon-lactone hydrolase | 0.047645 | 0.048087295 | 0.007734517 | 0.02568202 |
| K20608 | tetrahedral aminopeptidase | 0.047645 | 0.048091584 | 0.000710356 | 0.000166675 |
| K14188 | **D-alanine--poly(phosphoribitol) ligase subunit 2** | 0.047645 | 0.048095873 | 0.008249537 | 0.019178857 |
| K20487 | two-component system, OmpR family, lantibiotic biosynthesis sensor histidine kinase NisK/SpaK | 0.047645 | 0.048100163 | 0.006111161 | 0.021792519 |
| K00042 | 2-hydroxy-3-oxopropionate reductase | 0.047645 | 0.048104454 | 0.002712254 | 0.006392267 |
| K00637 | sterol O-acyltransferase | 0.047645 | 0.048108745 | 0.000749673 | 0.000398902 |

**Figure 11:**
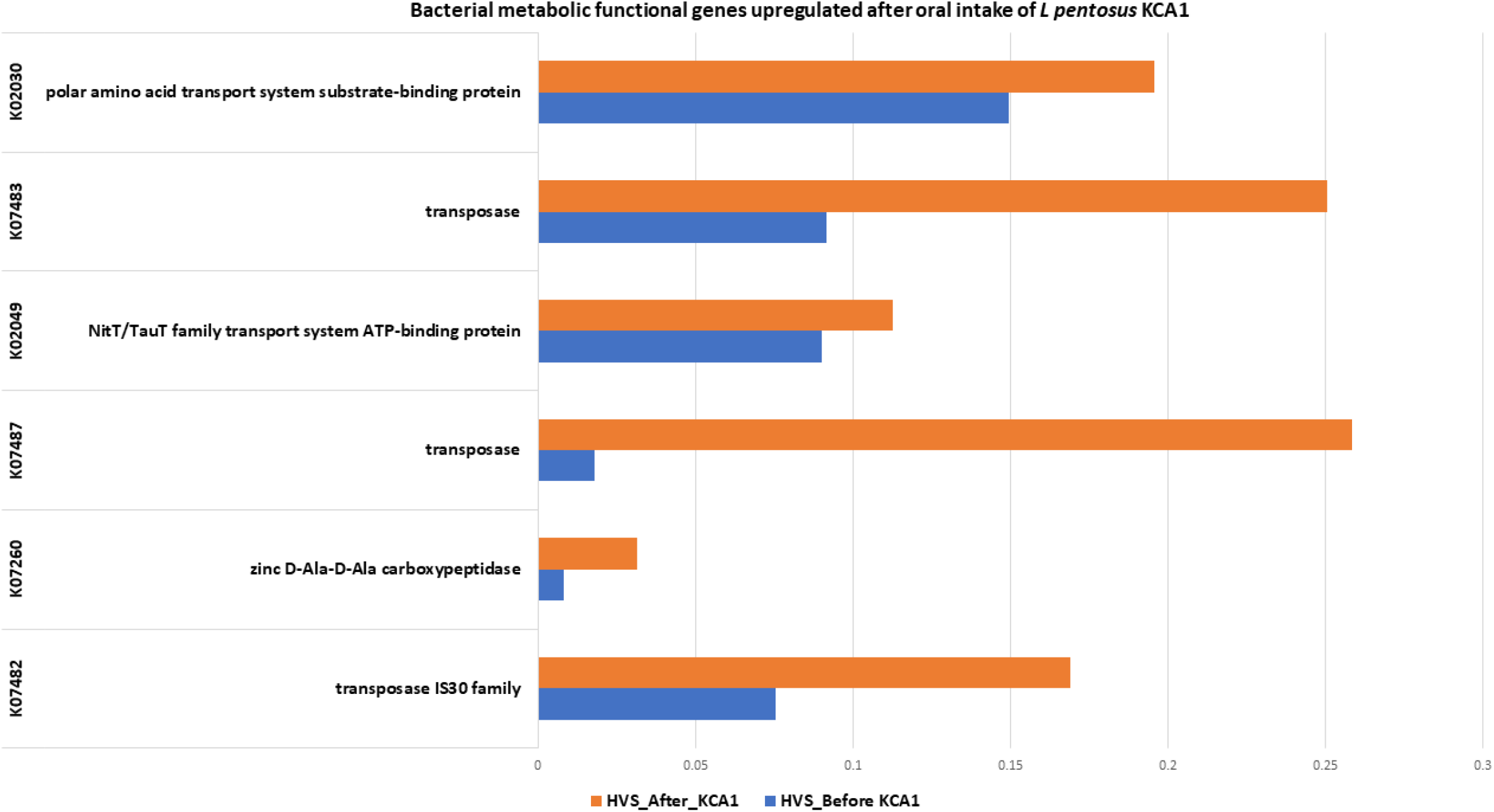
Bacterial metabolic functional genes upregulated in the vagina after oral intake of *L pentosus* KCA1.

Several bacterial metabolic functions in the gut were similarly affected. Interleukin 15 receptor alpha was downregulated significantly at P= 0.025347 and Methicillin resistance protein was also downregulated (P= 0.047645). Conversely, several microbial functional co-factors such as Menaquinone biosynthesis pathway, vitamin B6 (ko00750) metabolism, Biotin (ko00780) metabolism, Propanoate (ko00640) and Lipoic acid (ko00785) metabolisms were upregulated (**Figure 12**).

**Figure 12:**
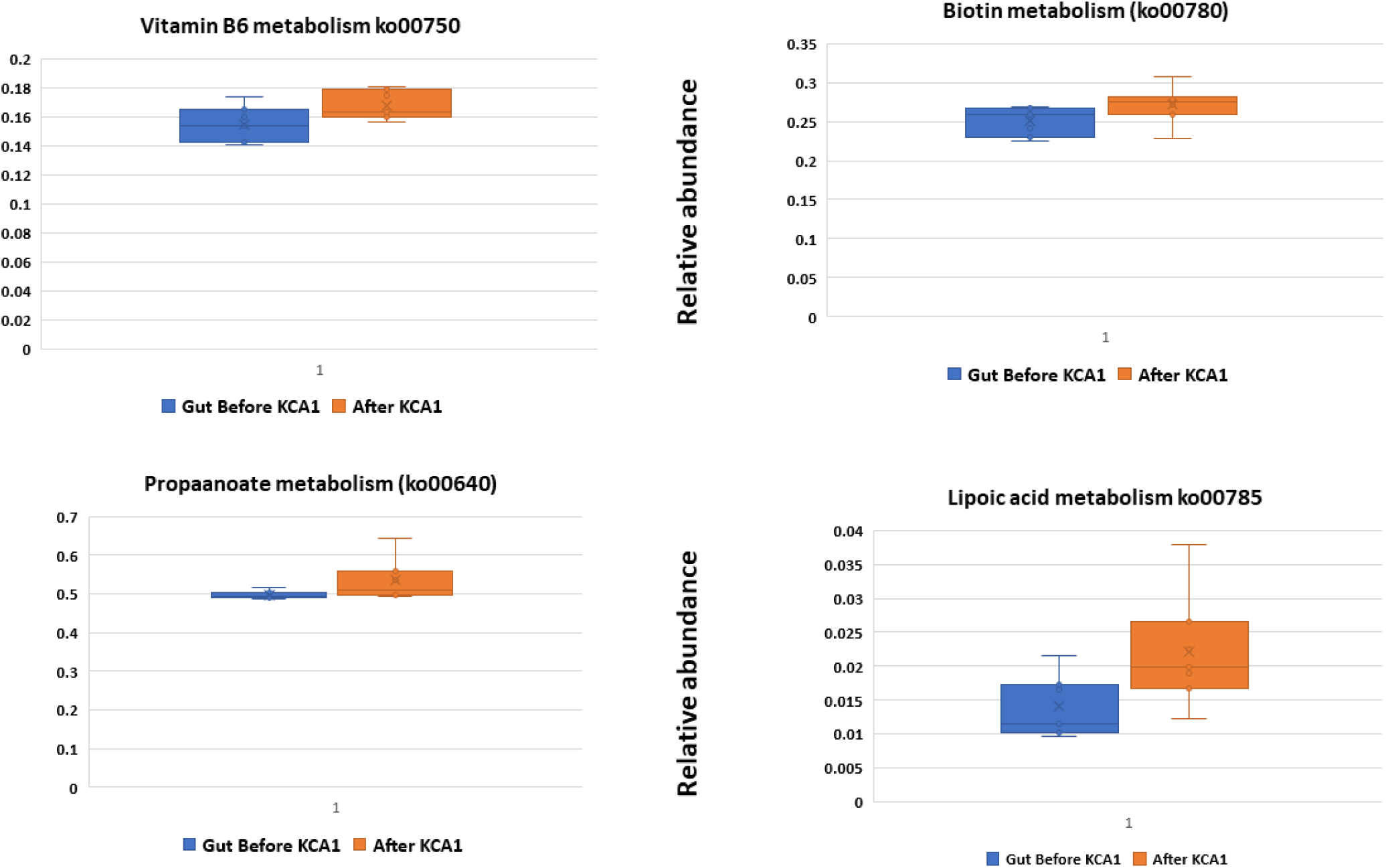
Bacterial metabolic functional genes upregulated in the gut after oral intake of *L pentosus* KCA1.

### Interleukin-1 beta (IL-1 beta)

There was a 2-fold down-regulation of IL-1 beta in contrast to IL-6 after consumption of KCA1 (**Figure 13**)

**Figure 13:**
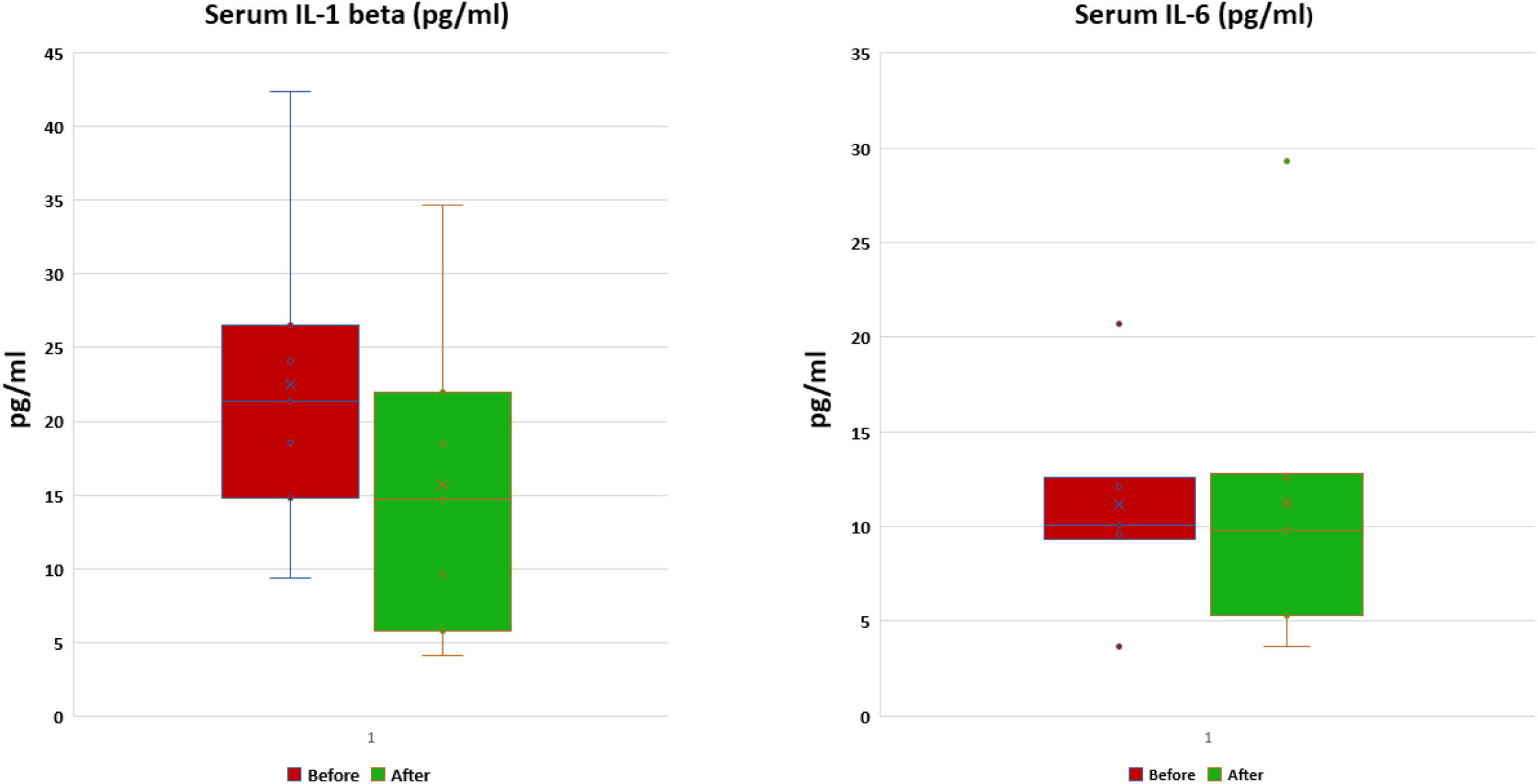
Showing box plot of the serum IL-1 beta and IL-6 before and after 14 days oral intake of *L pentosus* KCA1.

## DISCUSSIONS

In this study, we determined the composition of the vaginal and gut microbiota of women diagnosed with BV prior to and after 14 days consumption of *Lactobacillus pentosus* KCA1. In our previous study, we used metagenomics approach to decipher the composition of vaginal and gut microbiota of a small cohort of women diagnosed with BV. The study revealed the polymicrobial nature, phylogenetic diversity and species richness of the vagina and gut microbiome of reproductive age women diagnosed with BV.^**[5]**^ This present study is consistent with our previous study as the vaginal samples indicated fewer phyla in contrast to the gut. The consumption of KCA1 did not induce any significant difference in the species richness-alpha indices typified by ACE, CHAO1, Jackknife, and number of OTUs. However, at individual taxonomic categories, *L pentosus* KCA1 led to a decrease in the proportion of bacterial organisms associated with BV with a corresponding improvement in Nugent score typified by increase in Lactobacilli. It should be noted that in healthy women, the vagina is colonized predominantly by lactobacilli as we demonstrated in our previous study whereby the vagina of healthy, non-BV Nigerian women was colonized by Lactobacillus-dominated bacterial communities, in addition to the presence of other lactic acid–producing bacteria as revealed by 16S rRNA metagenomics.^**[21]**^ In a meta-analysis review, there was a statistically significant beneficial effect of probiotics observed in European populations after short-term follow-up days.^**[22]**^ The decrease of most of the bacteria associated with BV such as *Gardnerella vaginalis* in the vagina after consumption of KCA1 and the corresponding increase of the genera *Lactobacilli* suggest that KCA1 may be disrupting the bacterial biofilm associated with *Gardnerella vaginalis* survival.^**[23, 24]**^ *Lactobacillus pentosus* KCA1 is known to produce lactic acid and genes for the production of hydrogen peroxide and bacteriocins are encoded in the genome,^**[11]**^ of which several studies have shown to affect *G. vaginalis*.^[**25]**^ A recent study demonstrated the use of *Lactobacillus* strains against two different pathogens (*Gardnerella vaginalis* and *Atopobium vaginae*) responsible for bacterial vaginosis ^[**26]**^ Restoration of vaginal microbiota after insertion of a vaginal tablet have been reported.^[**27,28**]^ The decrease of the BV associated genera (*Porphyromonas, Prevotella, Gemella, Veillonella, Atopobium, Ureaplasma* and *Peptostreptococcus*) suggest that *Lactobacillus pentosus* KCA1 influences multi-genera metabolic activities. For example, a study reported certain strains of *Lactobacillus* in the vagina was found to suppress the epithelial inflammatory response to *Atopobium*, linked to bacterial vaginosis.^[**29]**^ The identification of some species not found before *L pentosus* KCA1 may indicate the possibility of cell-cross talk and quorum sensing thereby stimulating other related dormant cells to proliferate.^[**30]**^

While *Lactobacillus pentosus* KCA1 led to decrease in BV associated organisms in the vagina, it also decreased some BV associated organisms in the gut such as *Prevotella*. Conversely, it stimulated the proliferation of some bacteria known to be associated with health. For example, There was a 41.32% increase in *Faecalibacterium* especially, *Faecalibacterium prausnitzii*, which has been found to produce high amounts of butyrate and anti-inflammatory compounds.^[**31]**^ In the same vein, *Akkermansia muciniphila* proportion was elevated. It should be noted that *Akkermansia* is involved in metabolic functions and immune regulations and have been proposed as a promising probiotic.^[**32]**^

The *in silico* metabolic functional predictions indicated upregulation of some functions suggesting that *Lactobacillus pentosus* KCA1 impacted on the microbial activities. Intriguingly, the acquisition of microbial defense systems after consumption of KCA1 such as clustered regularly interspaced short palindromic repeats (CRISPR) is an indication that KCA1 can modulate other bacterial functions. The genome of *Lactobacillus pentosus* KCA1 encodes six CRISPR arrays with direct repeat (DR) length consensus exhibiting repeat polymorphisms. ^[**11**,**33]**^

The down-regulation of IL-15 by *Lactobacillus pentosus* KCA1 in the gut microbes is noteworthy, in that the cytokine correlates with adiposity and markers of the metabolic syndrome. Genome wide association (GWAS) have identified the gene encoding this receptor, IL15RA, which resides also on human chromosome 10p, a location linked to obesity and type-2 diabetes.^[**34]**^ The up-regulation of Menaquinone biosynthesis pathway and vitamin B6 vitamins suggest that consumption of *Lactobacillus pentosus* KCA1 has health attributes beyond the vagina. In addition to these attributes, KCA1 has direct effect on the human host as it down-regulated serum cytokine IL-1 beta, such suggesting modulation of inflammatory processes.

## CONCLUSION

Our findings suggest that *Lactobacillus pentosus* KCA1, taken orally, contributes positively to lowering pro-inflammatory cytokines, especially, IL-1 beta, with a significant decrease in the relative abundance of bacterial vaginosis-associated bacteria, and a corresponding increase of *Lactobacillus* genera. Increased metabolism of vitamin B6, biotin, lipoic acid and propanoate was observed in the gut. Bacterial genes related to defence systems were up-regulated in the vagina. The predicted metabolic functions indicate that some pathways in the vaginal microbial genes such as glycerol phosphoryl diester phosphodiesterase (K01126), mRNA interferase MazF (K07171), two-component system sensor histidine kinase, disease resistance protein (K13446), transposase IS30 family (K07482) were significantly up-regulated after consumption of *L pentosus* KCA1.

## Data Availability

All the relevant data generated in this study are included in the analysis

## ACKNOWLEDGMENT

We sincerely thank uBiome Inc, San Francisco, California, USA for awarding a grant-in-Kind to Dr. Kingsley Anukam in 2018 and for carrying out the metagenomics sequencing. We also thank Winclove Probiotics, Amsterdam, The Netherlands for Providing *Lactobacillus pentosus* KCA1 in powder form used for this study. Last but not the least, we thank all the women who participated in this study.

